# A survey of current anticoagulation patient education practices and development

**DOI:** 10.1101/2021.03.11.21252961

**Authors:** Aubrey E. Jones, John Saunders, Sara Vazquez, Angela Fagerlin, Daniel M. Witt

## Abstract

**Introduction:** Anticoagulants are high-risk medications widely used to prevent and treat thrombotic events, resulting in a need for adequate patient education to minimize harm. While anticoagulant patient education interventions can improve knowledge and surrogate outcomes, they may not represent current practice. Our objective was to determine provider perceptions of anticoagulation patient education at their institution.

**Methods:** A quantitative survey was distributed through a national professional organization and across several health systems. Questions included educational delivery methods, perceived effectiveness, evaluation of patient knowledge, development of patient education, and demographics.

**Results:** The 61 survey respondents were 79.0% female, 86.2% white, and an average age of 43. Most respondents (95%) complete patient education interventions in one session. Providers reviewed educational topics in 37% of daily patient interactions. 59% of respondents reported reasons for not performing patient knowledge checks included no formal process requiring knowledge checks, lack of time, and forgetting. The majority (93.4%) reported their patient education process was somewhat or very effective. The main reason underlying perceived less effective patient education was too much information in one session. Thirty-four respondents had some knowledge of patient education development at their institution. Most of these (82.9%) indicated their educational process’s development relied on expert opinion. In comparison, 22.9% indicated utilizing user-based design, and 10 (28.9%) respondents used learning theories.

**Conclusion:** While most providers felt patient education was effective, they reported reviewing education with patients daily. The lack of formal knowledge checks and best practices in developing patient education tools are significant gaps to address.

## Introduction

Anticoagulants are widely used to prevent and treat thromboembolic events, but are also high-risk medications.^1^ They are associated with increased risk of bleeding and are one of the top drugs causing emergency room visits in the United States.^2^ Taking anticoagulants incorrectly (i.e., non-adherence) can increase risk of bleeding or thromboembolic events. Drug interactions, and in the case of warfarin, dietary interactions also increase the risk of anticoagulant-related adverse events.^3^ Furthermore, warfarin is a narrow therapeutic index medication and requires regular blood test monitoring and dosing adjustments to ensure optimal anticoagulant effect.^3^ Thus, it is imperative that patients receive extensive high-quality patient education while taking anticoagulation therapy.

It has been shown that patients forget about 80% of what they learn in doctor’s offices, and of the 20% they do remember, only 50% is remembered correctly.^4, 5^ Currently the American Society of Hematology (ASH) guidelines for optimal management of anticoagulation therapy suggest supplemental patient education be provided to patients starting anticoagulation but provide no guidance regarding what that patient education should look like.^6^ Systematic reviews of patient education show that interventions of anticoagulant therapy patient education have ranged from intensive 90-120 minute training sessions to 5 minute educational videos.^7^ These studies have shown that most of these interventions do improve knowledge, but due to risk of bias, high levels of heterogeneity, and poor methodological quality, it is hard to know which supplemental educational methods are best.^7^ Additionally, developers of educational interventions rarely incorporate any learning theories or strategies when creating their tools or testing their effectiveness.^8^

Educational interventions reported in the literature likely represent only a small proportion of the supplemental anticoagulation therapy patient education provided on a routine basis. Provider views on patient education, barriers, and effectiveness have not been widely studied. One study assessing provider and patient views of anticoagulation in atrial fibrillation found that educational interventions were highly variable and often inadequate.^9^ Additionally, reported major barriers to providing patient education included lack of time and limited personnel.

The purpose of this study was to survey anticoagulation providers to determine provider attitudes and perceptions of supplemental anticoagulation therapy patient education delivery at their institution.

## Materials and methods

This quantitative study surveyed a national sample of anticoagulation providers. The survey was hosted in REDCap (an online, secure data collection tool) and distributed to potential participants via email. Invitations were sent to local anticoagulation clinics at the University of Utah Health, the Salt Lake City Veteran’s Affairs Medical Center, and Intermountain Health Care; and nationally to the Anticoagulation Forum list-serve, Veterans Affairs ambulatory care list-serve, and Kaiser Permanente’s anticoagulation provider distribution list. A hyperlink to the survey was included in the recruitment email.

The recruitment email contained a background explanation of the survey purpose as well as consent language. Consent was obtained electronically upon entering the survey. Providers who received the recruitment email had 12 weeks to complete the survey with two reminder emails sent at 4 and 8 weeks before the survey closed. Providers were able to save their answers and return to the survey at a later time if needed.

Any health care provider who provides anticoagulation patient education, including physicians, pharmacists, mid-level providers (physicians assistants and nurse practitioners), nurses, licensed practical nurses (LPNs), and medical assistants (MAs) was eligible to complete the survey. Providers were excluded if they were under the age of 18 years or did not speak English.

### Survey

Survey questions included basic demographic information including age, gender, self-identified race and ethnicity, and practitioner type (see above). Subsequent survey questions were aimed at ascertaining types of supplemental education delivery used, details regarding site-specific educational processes, perceived effectiveness of and satisfaction with current delivery of supplemental education, and methods used to create patient education. Branching logic was used to gather further details about reasons for perceived ineffective/inefficient education, evaluation of education processes, checking for patient understanding, and supplemental patient education materials. The complete survey is provided in Appendix A. Survey questions included multiple choice questions, free text responses, and Likert scales.

### Analysis

Survey responses were summarized using basic descriptive statistics such as proportions for categorical variables and means and standard deviation for continuous variables. Likert scale responses were dichotomized to simplify reporting.

## Results

In total 61 providers completed the survey. Due to the use of multiple list-serves and group emails, we were unable to calculate a response rate. The average age of respondents was 43.3 years (Standard Deviation (SD) 11) and 47 (78.3%) were female (Table 1).

**Table 1.** Characteristics of Survey Respondents

|  | N, % |
| --- | --- |
| Age, mean, (standard deviation) | 43, (11) |
| Female | 46, 78% |
| White race | 50, 86.2% |
| Non-Hispanic ethnicity | 55, 93.2% |
| Provider type |  |
| Nurse practitioner | 2, 3.3% |
| Registered nurse | 8, 13.3% |
| Physician | 2, 3.3% |
| Pharmacist | 48, 80% |

Most respondents were white (86.2%) and of non-Hispanic ethnicity (93.2%). The majority (80.3%) were pharmacists, with registered nurses (13.1%), physicians and nurse practitioners (3.3% each) making up the rest of respondents. 57.4% of respondents indicated that they were involved in the creation or selection of supplemental patient educational materials.

### Nature of Patient Education

The majority of respondents indicated that all new patient education is completed in one session (95.1%) (Figure 1a). Educational sessions last on average 35 minutes (SD 15.2) for warfarin and 22.5 minutes (SD 13.2) for DOACs (Figure 1b). Of providers indicating that more than one patient education sessions were used, average duration was 56 minutes (SD 30.6) for warfarin and 45 mins (SD 14.4) for DOACs per session. All providers used printed supplemental education materials. Other types of education materials included videos (14.8%), computer-based (8.2%), group classes (6.6%), and other material types (3.3%) (Figure 2).

**Figure 1.**
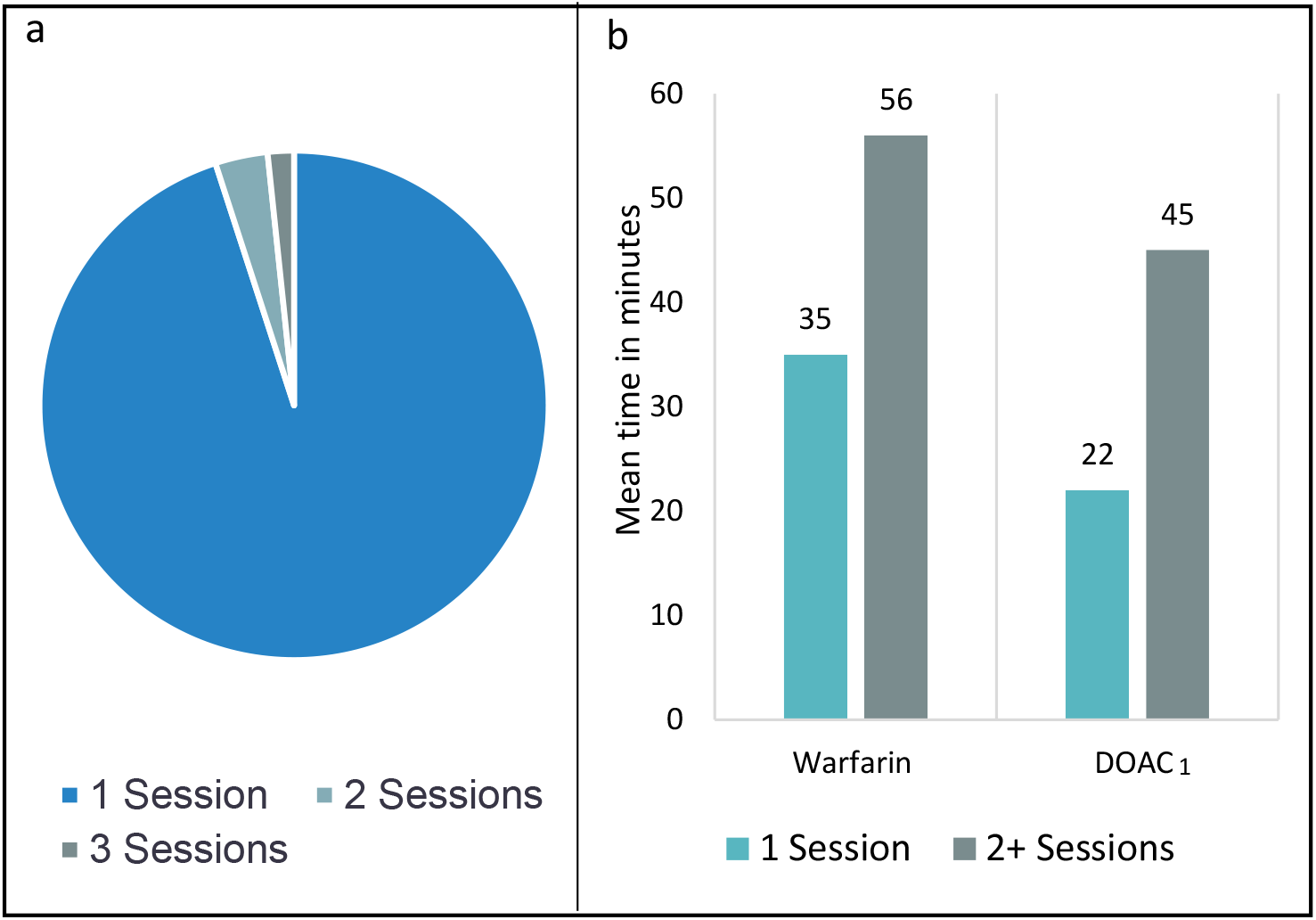
**Figure 1a:** Reported number of sessions it takes providers to complete new patient education session; **Figure 1b:** Average length of new patient education for warfarin and direct oral anticoagulants (DOAC) per session classified by number of sessions taken to complete

**Figure 2.**
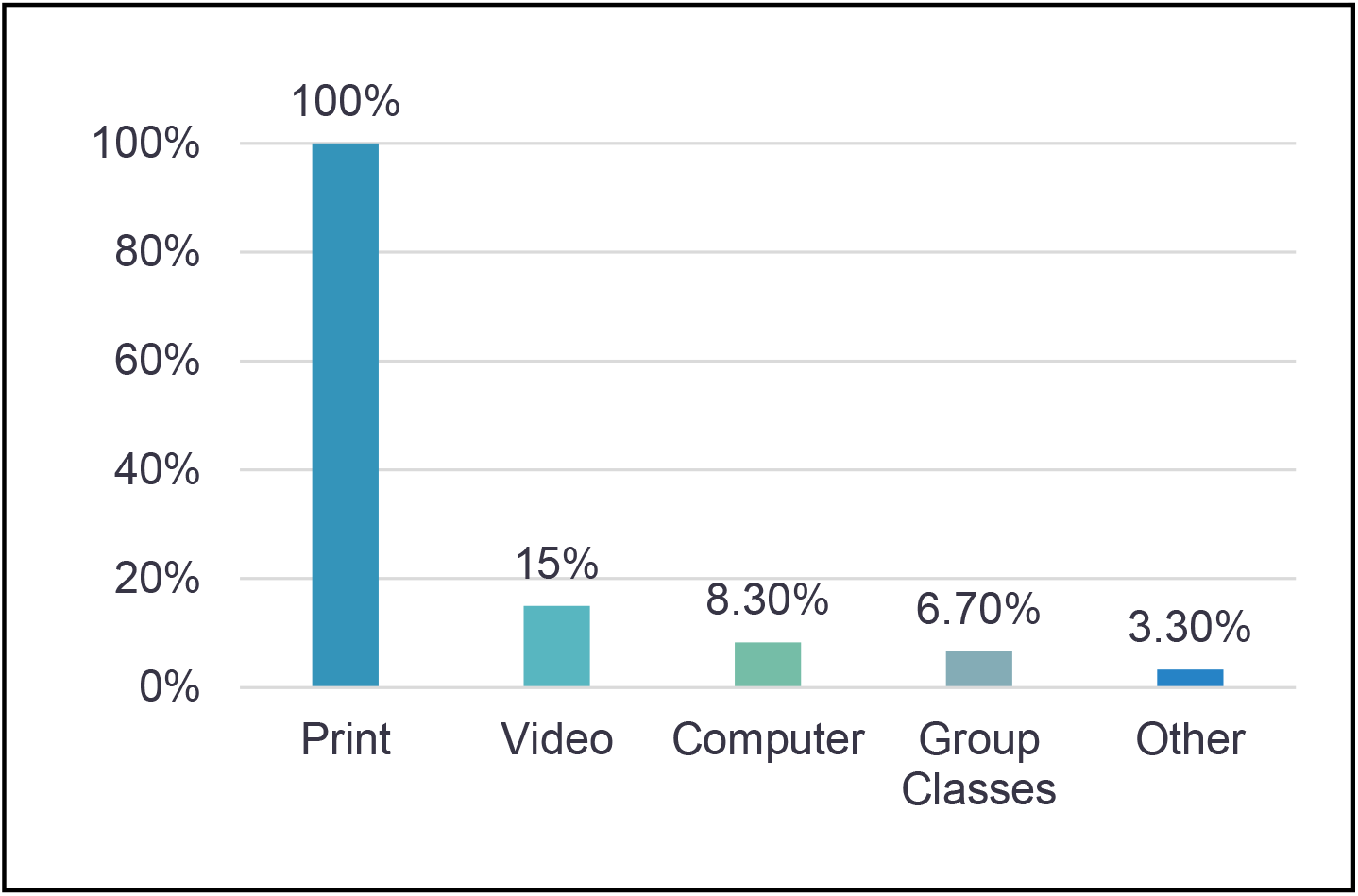
Types of Supplemental Education used by anticoagulation providers for warfarin and DOAC new patient

### Assessing Patient Knowledge

We asked respondents whether they used formal knowledge checks to ensure patient understanding – such as written quizzes or the teach-back method, versus less standardized informal knowledge checks.

27.9% indicated using both formal and informal knowledge checks, (Table 2), 4.9% just did a formal check, 57.4% did just informal checks, and 8.1% didn’t do any type of knowledge checks. Of those performing formal knowledge checks who responded to the question (n=19), 6 used a combination of the following types of tests, just 1 respondent (5.2%) used only a written test, 57.9% just used the teach-back method, and 5.2% just did a verbal test. One of the respondents who utilized multiple test types used the Indian Health Services model of asking “What did your provider tell you this medication was for?”, “How were you told to take it?”, and “What were you told to expect?”.^10^ Stated reasons for performing knowledge checks included patient demonstrated a deficit of knowledge (89.7%), the provider felt like the patient did not understand (77.6%), and it was a required part of the educational process (20.7%). Only 2 (5.6%) respondents reported feeling that knowledge checks were not useful. On average, respondents reported reviewing some aspect of patient education in 37% of routine daily patient interactions. The most common topics reviewed included how dietary vitamin K affects the INR (78.7%), the importance of notifying anticoagulation providers of new medications (55.7%), compliance or the importance of not missing doses (41.0%), and what the INR means (37.7%), (Figure 3).

**Table 2.** Completion of knowledge checks by anticoagulation providers

| **Types of checks, n=61** | **N, %** |
| --- | --- |
| Both Formal and Informal Checks | 20, 27.9% |
| Formal Check Only | 3, 4.9% |
| Informal Check Only | 35, 57.4% |
| No check | 6, 9.8% |
| **Types of knowledge check, n=19<sup>1</sup>** |  |
| Multiple types of tests<sup>2</sup> | 6, 31.6% |
| Written Test | 1, 5.2% |
| Teach Back | 11, 57.9% |
| Verbal Test | 1, 5.2% |
| **When do you use knowledge checks, n=57<sup>1</sup>** |  |
| Formal part of education process | 12, 20.7% |
| If I think patient does not understand | 45, 77.6% |
| Patient behavior demonstrates deficit of knowledge | 52, 89.7% |
| **Why are knowledge checks not used, n=36<sup>1</sup>** |  |
| Take too much time | 11, 30.6% |
| Not useful | 2, 5.6% |
| I forget to use them | 7, 19.4% |
| Not part of our formal process | 18, 50% |
| I never learned to use them | 1, 2.8% |
| Other | 7, 19.4% |

<sup>1</sup>Respondents could give more than response
<sup>2</sup>Types of tests included any combination of written, teach back method, the Indian Health Services model, and verbal tests

**Figure 3.**
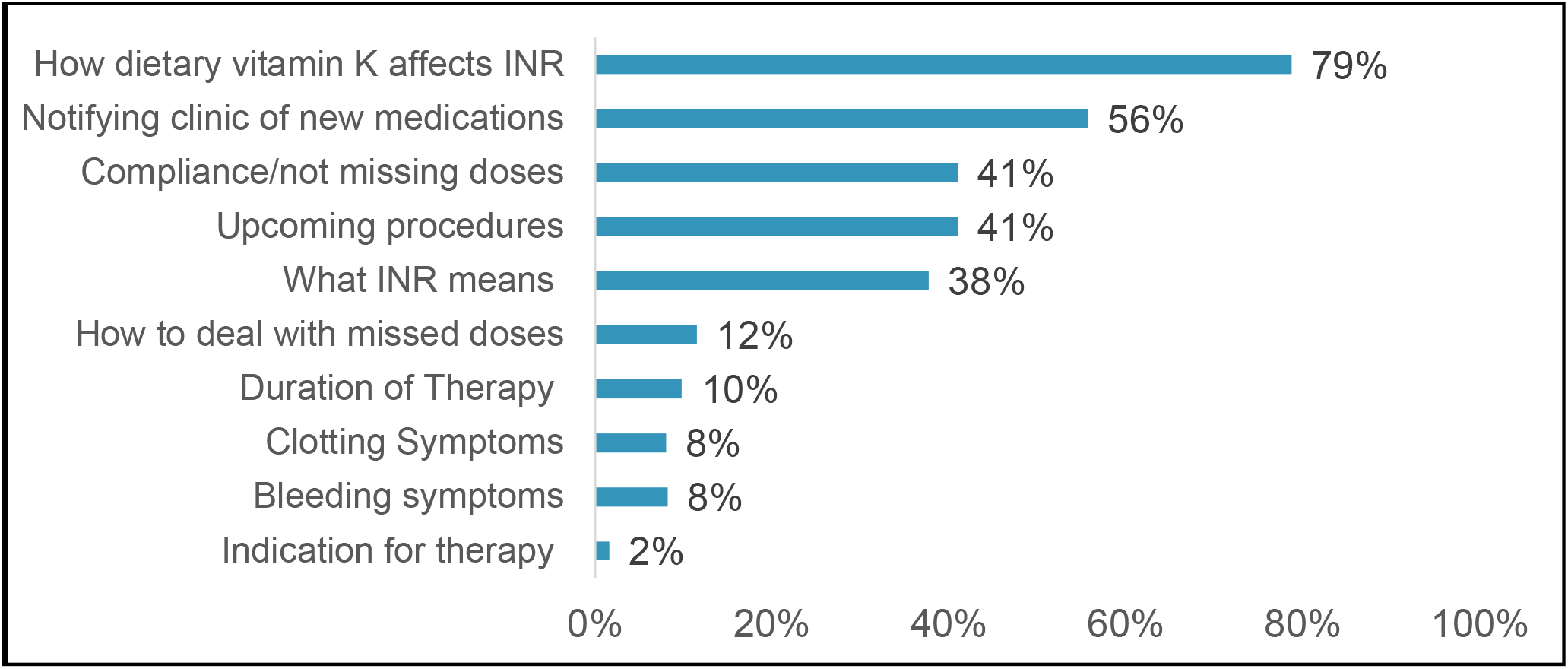
Most common topics providers spend time reviewing with patients

### Assessment of Patient Education

The majority of respondents reported their education was somewhat or very effective (93.4%). In a free text response asking the top 2-3 reasons they felt their education strategies were not as effective as they could be (n=43) the most common theme reported was too much information to cover in one session (41.8%). Other less commonly reported themes included patient apathy or lack of engagement in learning, verbal only education, providing education over the telephone, patient cognition or hearing/visual impairment, and lack of formal follow-up to assess patient understanding. Fewer respondents rated their education process as being efficient (83.6%). Only 10 (16.4%) respondents reported that the patient education processes at their institution has been formally evaluated. Of these, two respondents found that the current process was meeting their needs, and 7 reported it mostly met their needs but some changes were necessary, with one not responding.

### Creation of Patient Education

Just over half (57.4%) of respondents reported involvement in or knowledge of the process for creating patient education materials at their institution. Regarding the process for determining what to include in the patient education process, the majority relied on expert opinion (29, 82.9%), while only 22.9% included patients in developing the process (user-based design) (Figure 4). Ten respondents (29.4%) reported using validated learning theories in their educational materials while 42.9% (n=15) and 28.6% (n=10) stated they did not incorporate learning theories or didn’t know, respectively. Only six respondents (17.6%) reported doing any pilot testing of their educational process, but 27 (77.1%) reported following existing guidelines or frameworks when creating their educational materials.

**Figure 4.**
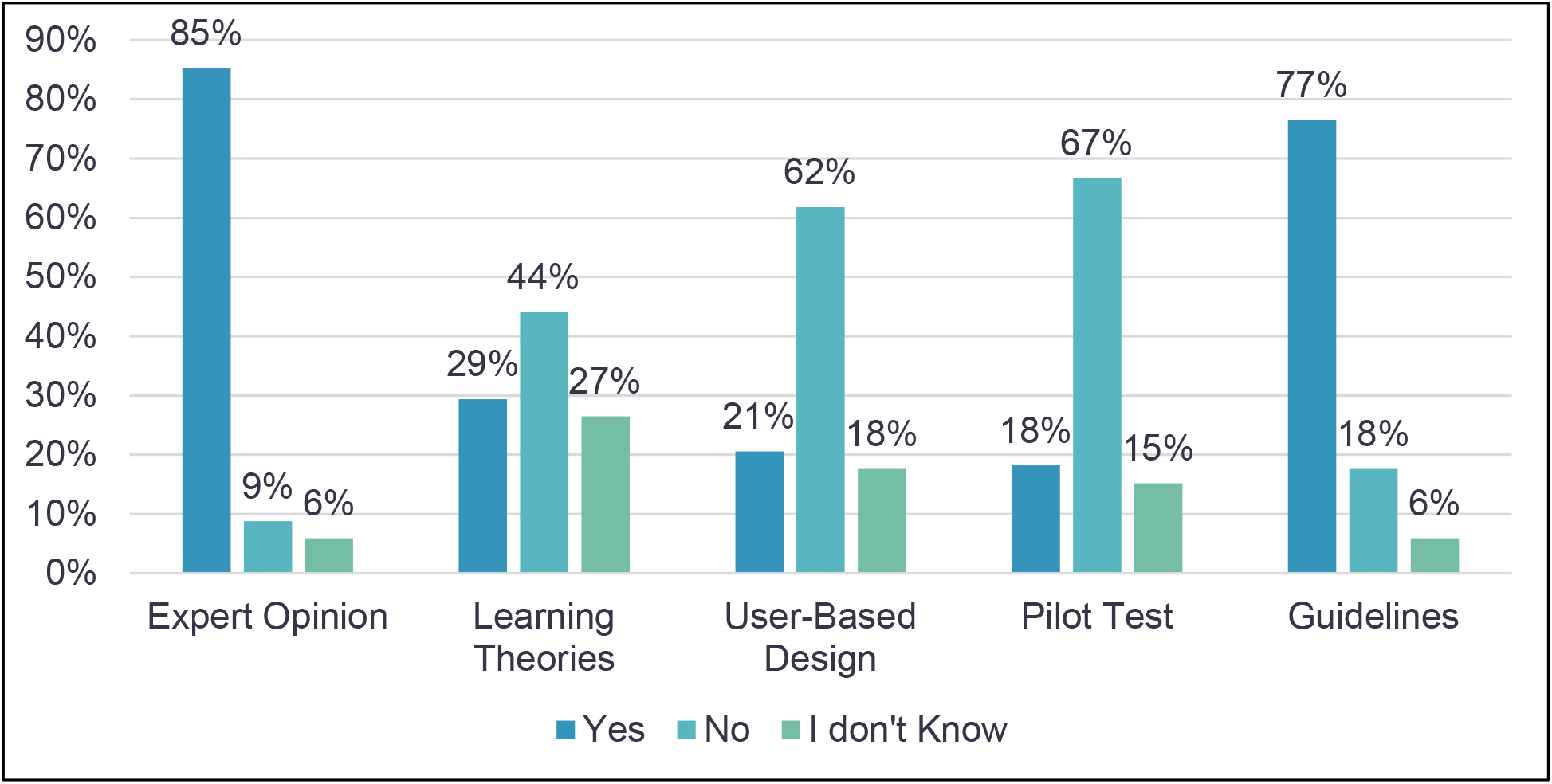
Different methods of patient education development that are used by providers involved in creating anticoagulation patient education (n=34)

## Discussion

Our findings demonstrate that while the majority of providers feel their education is at least somewhat effective, they are still reviewing educational points during about 40% of their daily patient interactions. Many providers mentioned that a lack of time and overwhelming patients with too much information at once were the main reasons they felt their current educational process was less effective. This is consistent with studies that have shown that education is more likely to be retained when spread out over multiple sessions and learners are given opportunities for information retrieval (e.g., quizzes, teach-back method).^11–14^ The observation that dietary issues with warfarin is a common issue needing to be readdressed is consistent with previous patient knowledge assessment studies.^15^

It is important to point out that very few respondents in our study indicated that the effectiveness of their educational process had ever been formally evaluated, and of those who had, many found improvement was needed. Additionally, the minority of providers indicated that formal knowledge checks were incorporated into their patient education process, and several providers reported feeling that lack of formal knowledge checks was a reason underlying why their education process was less effective. Since most providers reported that they do not perform knowledge checks because of forgetfulness, lack of time, or lack a formal incorporation into the educational process, this is a potential target for interventions aimed at improving patient education processes. Formally assessing patient knowledge, and testing learners’ knowledge has been shown to increase retention and improve learning.^16–18^

Few providers are following best practice recommendations of incorporating patients into the creation of educational materials, pilot-testing, or incorporating validated learning theories. This observation highlights significant improvement opportunities in developing effective patient education materials. While respondents reported that they followed guidelines when creating patient education materials, it is possible they were referring to clinical guidelines that suggest providing supplemental patient education for anticoagulated patients as opposed to educational guidelines as the question did not specify which type of guidelines were being referred to.

Optimal patient education development should incorporate learning theories and strategies. Clinical education that has implemented them into their development have shown improved clinical outcomes. However, a systematic review of thrombosis and hemostasis education found that only two studies (one in pediatrics, one in adults) out of 16 incorporated learning theories into their patient education.^8^ Those that did incorporate learning theories in their development didn’t measure outcomes that would lead to conclusions about whether learning based education is better or worse. They point out that as learning strategies arise from different learning theories, use of these theories when evaluating different situations would lead to more effective strategies being chosen for that patient education. The review recommends conducting studies designed to compare theory-based patient education to standard of care in order to overcome the currently weak evidence that exists for determining optimal patient education.

The lack of learning theories or strategies being incorporated into patient education interventions could be do to several things. There are no formal regulations in clinical guidelines on how patient education should be designed, or where some guidance does exist, clinicians may not be aware. Clinicians who manage services are often those who end up formulating education materials or programs that will work in their system. Often these people are never formally trained on what learning theories or strategies are, much less how to implement them in materials or programs or evaluate them. This lack of guidance and knowledge on how to create patient education calls for

A limitation of our study is that providers were not asked to specify their institution, so multiple providers may have reported their experience with the same patient education materials. While this may not have influenced how providers perceive the effectiveness of, or their own personal application of educational materials, if multiple respondents were involved in creating patient education materials at a given site, answers to some question may have been duplicated. One major limitation in our survey was that we failed to ask providers was about whether they deliver patient education face to face or over the phone. This may affect effectiveness of patient education as visual cues can be helpful when discussing topics and perceiving confusion or understanding. Although the survey was sent to several large national anticoagulation provider list serves, only 61 responses were included in the analysis. We were unable to calculate a survey response rate as the true denominator value of providers who received the invitation email is unknown due to the use of multiple list-serves but the sample was smaller than anticipated given previous studies using this same methodology.^19^ As there wasn’t huge variation in our responses, it could be that non-responders wouldn’t change our findings much rather than if we had seen a huge variation in responses.. However, survey respondents may have been individuals more likely to respond because they are involved or interested in patient education, and thus may be unusual compared to non-responders.

## Conclusion

While most providers rated their patient education processes for new patients receiving anticoagulation therapy as effective, many daily patient interactions are still spent reviewing educational information with patients. The lack of formal knowledge checks in the patient education process, and sufficient time are potential major gaps contributing to less effective patient education. Additionally, the majority of patient education development does not follow best practice recommendations. Our results indicate a need for a more standardized guidance for creating and carrying out anticoagulation therapy patient education based on validated educational theories.

## Data Availability

The authors confirm that the data supporting the findings of this study are available within the article [and/or] its supplementary materials.

## References

1. Ageno W, Gallus AS, Wittkowsky A, Crowther M, Hylek EM, Palareti G. Oral anticoagulant therapy: Antithrombotic Therapy and Prevention of Thrombosis, 9th ed: American College of Chest Physicians Evidence-Based Clinical Practice Guidelines. Chest. Feb 2012;141(2 Suppl):e44S–e88S. doi:10.1378/chest.11-2292

2. Budnitz DS, Shehab N, Kegler SR, Richards CL. Medication use leading to emergency department visits for adverse drug events in older adults. Annals of internal medicine. Dec 4 2007;147(11):755–65.

3. Witt DM, Clark NP, Kaatz S, Schnurr T, Ansell JE. Guidance for the practical management of warfarin therapy in the treatment of venous thromboembolism. Journal of thrombosis and thrombolysis. Jan 2016;41(1):187–205. doi:10.1007/s11239-015-1319-y

4. Anderson JL, Dodman S, Kopelman M, Fleming A. Patient information recall in a rheumatology clinic. Rheumatol Rehabil. Feb 1979;18(1):18–22.

5. Kessels RP. Patients’ memory for medical information. J R Soc Med. May 2003;96(5):219–22. doi:10.1258/jrsm.96.5.219

6. Witt DM, Nieuwlaat R, Clark NP, et al. American Society of Hematology 2018 guidelines for management of venous thromboembolism: optimal management of anticoagulation therapy. Blood Adv. Nov 27 2018;2(22):3257–3291. doi:10.1182/bloodadvances.2018024893

7. Paquette M, Witt DM, Holbrook A, et al. A systematic review and meta-analysis of supplemental education in patients treated with oral anticoagulation. Blood Adv. May 2019;3(10):1638–1646. doi:10.1182/bloodadvances.2019000067

8. Hews-Girard J, Guelcher C, Meldau J, McDonald E, Newall F. Principles and theory guiding development and delivery of patient education in disorders of thrombosis and hemostasis: Reviewing the current literature. Res Pract Thromb Haemost. Oct 2017;1(2):162–171. doi:10.1002/rth2.12030

9. Borg Xuereb C, Shaw RL, Lane DA. Patients’ and health professionals’ views and experiences of atrial fibrillation and oral-anticoagulant therapy: a qualitative meta-synthesis. Patient Educ Couns. Aug 2012;88(2):330–7. doi:10.1016/j.pec.2012.05.011

10. Lam N, Muravez SN, Boyce RW. A comparison of the Indian Health Service counseling technique with traditional, lecture-style counseling. J Am Pharm Assoc (2003). Sep-Oct 2015;55(5):503–10. doi:10.1331/JAPhA.2015.14093

11. McDaniel M. Put the SPRINT in knowledge training: Training with SPacing, Retrieval, and INTerleaving. 2012;

12. Birnbaum MS, Kornell N, Bjork EL, Bjork RA. Why interleaving enhances inductive learning: The roles of discrimination and retrieval. Memory & cognition. 2013;41(3):392–402.

13. Finn B, Roediger III HL. Enhancing retention through reconsolidation: Negative emotional arousal following retrieval enhances later recall. Psychological science. 2011;22(6):781–786.

14. Pan SC, Rickard TC. Transfer of test-enhanced learning: Meta-analytic review and synthesis. Psychol Bull. 07 2018;144(7):710–756. doi:10.1037/bul0000151

15. Wang Y, Kong MC, Lee LH, Ng HJ, Ko Y. Knowledge, satisfaction, and concerns regarding warfarin therapy and their association with warfarin adherence and anticoagulation control. Thrombosis research. Apr 2014;133(4):550–4. doi:10.1016/j.thromres.2014.01.002

16. Roediger III HL, Karpicke JD. The power of testing memory: Basic research and implications for educational practice. Perspectives on psychological science. 2006;1(3):181–210.

17. Roediger III HL, Putnam AL, Smith MA. Ten benefits of testing and their applications to educational practice. Psychology of learning and motivation. Elsevier; 2011:1–36.

18. Barcellona D, Contu P, Marongiu F. Patient education and oral anticoagulant therapy. Haematologica. Oct 2002;87(10):1081–6.

19. Gardner T, Vazquez SR, Kim K, Jones AE, Witt DM. Providers’ utilization and perceptions of warfarin dosing algorithms. Thrombosis research. Sep 2019;183:4–12. doi:10.1016/j.thromres.2019.09.002

